# Association of Pre-COVID-19 Lymphocytopenia with Fatality

**DOI:** 10.1101/2020.10.02.20200931

**Authors:** W.R. Burack, P. Rock, D. Burton, X. Cai

**Affiliations:** Pathology and Laboratory Medicine, University of Rochester Medical Center; Biostatistics and Computational Biology, University of Rochester Medical Center

## Abstract

Lymphocytopenia during the COVID-19 has been associated with fatality. We tested whether pre-existing lymphocytopenia reported prior to any possible exposure to SARS-COV2 (from 2010 to 2019) was associated with fatality. Using all patients diagnosed on testing in a single regional laboratory, we identified 1137 subjects with PCR positive for SARS-COV2 and at least one available complete blood count from the decade prior to any possible exposure to the virus. Bivariate analysis indicated an association between pre-existing lymphocytopenia (defined as absolute lymphocyte count <0.9×10^9^ /L) and fatality (18% versus 4%). Furthermore, a logistic regression model, accounting for both patient age and number of blood counts obtained, indicated the subjects with pre-existing lymphocytopenia were 1.4 times as likely to die. Because the absolute lymphocyte count is almost universally available and easily interpreted, this biomarker of the risk of fatality could be widely useful.

## INTRODUCTION

The clinical course of COVID-19 is remarkably variable. For example, while 20% of SARS-CoV2 positive patients in their eighth decade succumb to COVID-19, at least a similar fraction remain asymptomatic. This observation has motivated a number of efforts to identify prognostic markers that pre-exist Covid, such as co-morbidities or genotypic features, particularly those that may be reasonably associated with respiratory failure or other common features of the disease. Comorbidities are a clear risk factor for fatality and recently several germline genetic features have been associated with disease severity. However, there is not a simple-to-understand and widely available baseline laboratory value which can stratify risk.

Lymphocytopenia is a common feature of COVID-19 and, critically, further decreases in the absolute lymphocyte count (ALC) in patients with COVID-19 portend respiratory failure and death^1-5^. Given this association between fatality and lymphocytopenia *during* COVID-19, we hypothesized that lymphocytopenia *prior* to any possible infection by the virus could be a risk factor for fatality.

## METHODS

To test for an association between pre-existing lymphocytopenia and fatality, we sought subjects who were PCR-positive for SARS-CoV2 and with complete blood counts (CBC) from 2019 or earlier, thus ensuring that these complete blood counts pre-dated any potential exposure to SARS-CoV2 by at least 3 months. After data were collected, descriptive statistics were performed when means and standard deviations were presented for continuous data, and frequencies were presented for categorical data. Bivariate associations between patient death status and their pre-Covid19ALC as well as patient demographic categories were examined, in which chi-square tests were applied. A Logistic regression model of patient death status was further conducted against patient pre-Covid19ALC, its testing frequencies, and patient age.

## RESULTS

Of 1288 adults subjects with PCR-positive COVID-19 nasopharyngeal swabs in our medical system (3/15-5/22/2020), 1137 (median age 61) had ≥1 CBC during the decade preceding possible exposure to SARS-CoV2 (median 4.5; 2010-12/1/2019) and 617 had also had ≥1 CBC in 2020. Subjects were followed to 6/1/2020. For the 1137 subjects with ALCs obtained prior to COVID-19, the median pre-Covid19 ALC was 1.7×10^9^/L, and 57 (5%) had pre-Covid19 lymphocytopenia (defined as median ALC <0.9×10^9^/L). 71 deaths were observed (6.2%).

As expected, the case-fatality rate was associated with lymphocytopenia in 2020 (18% for subjects with lymphocytopenia (52/282) vs 4% for all others (15/335); P< 0.0001 Chi-square). Also consistent with previous reports, fatality was age-dependent (18-49: 0.7% (3/425); 50-64: 2.9% (6/208); 65-79:9.2% (25/271); 80+:16% (37/233); *χ*^2^=67, P<.0001).

Bivariate analyses showed association of fatality with pre-existing lymphocytopenia (10/57 [17.5%] vs. 61/1080 [5.1%]; Fisher exact test P<0.0001). Among 617 subjects with an ALC both prior to 2020 and during COVID19 treatment, 282 [46%] were lymphocytopenic in 2020; lymphocytopenia in 2020 was associated with pre-existing lymphocytopenia (30/252 [10%] vs 5/330 [1.5%]; *χ*2=23, P<.0001; table 1; figure 1) and with fatality (52/282 [18%] vs 15/335 [4%], table 2).

**Table 1.**
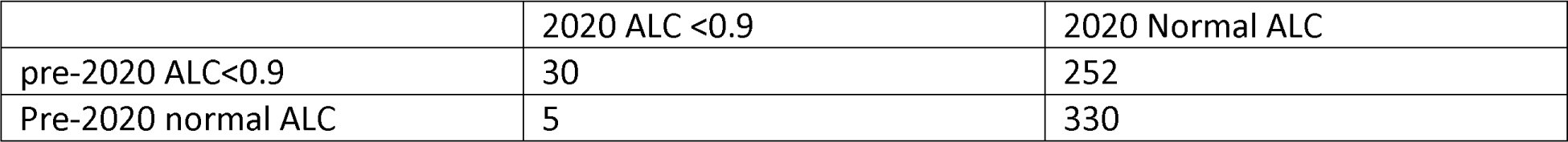

**Table 2.**
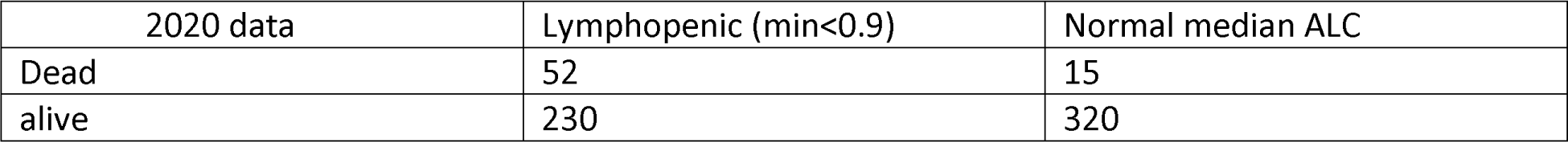

**Figure 1.**
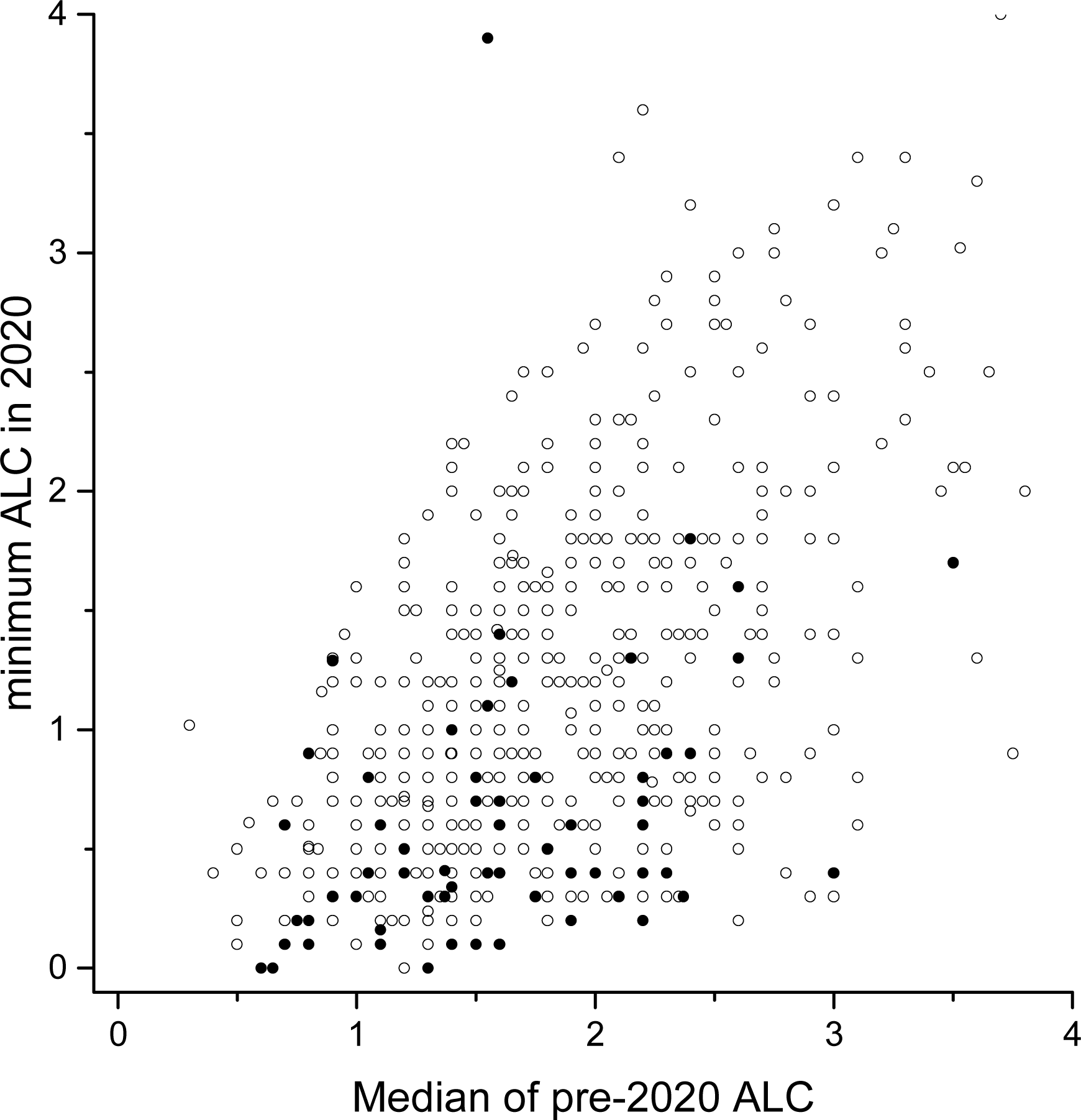
The relationship between median pre-2020 ALC and the minimum ALC reported in 2020 for COVID positive patients. Filled symbols are deceased subjects.

A logistic regression model, accounting for patient age and the number of ALCs prior to 2020 (a surrogate for the extent of interaction with the medical system), showed that patients with pre-existing lymphocytopenia were 1.4 times more likely to die compared to those without pre-existing lymphocytopenia (table 3; P=0.023).

**Table 3.**

| Table 3 | Analysis of Maximum Likelihood |  |  |  | Odds Ratio Estimates |  |  |
| --- | --- | --- | --- | --- | --- | --- | --- |
| Parameter | Estimate | Standard Error | Wald Chi-Square | Pr > ChiSq | Point Estimate | 95% Wald Confidence Limits |  |
| Intercept | -4.9 | 0.61 | 66 | <.0001 |  |  |  |
| Pre-20202 ALC < 0.9 vs ≥ 0.9 | 0.88 | 0.39 | 5.1 | 0.023 | 2.4 | 1.1 | 5 |
| Age 50-64 vs 18-49 | 1.48 | 0.72 | 4.0 | 0.046 | 4.2 | 1.0 | 17 |
| Age 65-79 vs 18-49 | 2.68 | 0.62 | 18 | <.0001 | 14 | 4.1 | 47 |
| Age 80+ vs 18-49 | 3.28 | 0.62 | 28 | <.0001 | 26 | 7.6 | 87 |
| Number of CBCs <3 vs ≥ 3 | -0.096 | 0.33 | 0.083 | 0.77 | 0.91 | 0.47 | 1.7 |

## DISCUSSION

These data show that pre-2020 lymphocytopenia is associated with an increased odds ratio of death. On review of the pre-2020 medical records, no features overtly distinguished those patients with lymphocytopenia from those without. Among the 10 deceased subjects with pre-COVID lymphocytopenia, the pre-2020 ALCs were only moderately reduced (range 0.6-0.8; median 0.725). While very large data sets would be required to define the performance of any specific cutpoint, these data suggest that a population enriched in particularly vulnerable subjects may be operationally identified as having an ALC <0.9×10^9^/L.

The mechanisms by which a decreased pre-Covid ALC may affect fatality are unclear. There is widespread interest in whether baseline immune-status affects fatality and simply the lymphocyte number may be a reflection of the immune status. Alternatively, reduced ALC may reflect therapies for various disorders that would require competent review of each medical record to identify. Because the ALC is a simple and almost universally available test, the association of pre-Covid lymphocytopenia with fatality may allow individuals and providers to succinctly communicate personal risk, facilities to streamline the triage of resources for isolation, and epidemiologists to improve pandemic-scale modeling.

## Data Availability

available from the corresponding author

## Contributions

WRB: conceived the work, wrote the manuscript, conducted research, and analyzed data; PJR: conducted research; DB: analyzed data; XC: analyzed data

